# More than the sum of its parts? An analysis of the factors impacting the content of antenatal care consultations in the Democratic Republic of Congo

**DOI:** 10.1101/2025.01.17.25320735

**Authors:** Rui Han, Brittany Hagedorn

## Abstract

**Background:** Previous studies have shown that in lower and middle-income countries, although antenatal care (ANC) coverage has increased significantly, maternal mortality remained high due to poor quality of care. Despite established protocols, recommended actions are not consistently completed and the causes are not well understood. We sought to unpack this by analyzing direct observations of ANC consultations from the Democratic Republic of Congo (DRC).

**Methods:** We conducted secondary analysis of data from the Health System Strengthening for Better Maternal and Child Health Results Project (2015-2022) evaluation. We examined completion rates of twenty recommended actions for a first ANC visit. We identified contributing factors using chi-squared tests on completion rates, in samples stratified by availability of essential items and provider characteristics. We used regressions to estimate the relative importance of facility, provider, and patient characteristics in determining completion rates and the effect of availability of the essential item for different care components.

**Results:** We observed a statistically significant increase between baseline and endline in supply-side readiness (equipment and supplies) and quality of care for six recommended actions. While structural readiness was a significant predictor of quality, we found that provider qualifications and female gender had significant positive associations with completion rates, partially explaining the observed variation in care quality. For example, female A1 nurses prescribed tetanus vaccination for 70% of patients, male A1 nurses to 55%, and midwives to 51%. After decomposing the reasons for poor quality, we found that in addition to supply-side gaps (64% at baseline, 51% at endline), the know-do gap contributed to 30% of incompletions at baseline (21% at endline). Patient characteristics were rarely predictive of quality.

**Conclusions:** While supply-side gaps continue to be important barriers to service quality for ANC visits, especially preventive treatments and diagnostics, addressing these alone is inadequate. This is especially true for the physical exam, where interpersonal and behavioral barriers persist and we see evidence of gendered differences in quality of care. Interventions to improve the performance of all providers is critical to ensure that every person receives the best possible care.

## Background

Ensuring adequate quality of care is a pressing issue in lower and middle-income countries (LMICs), where health systems often face multiple challenges such as under-equipped facilities, inadequate training of the workforce, and weak governance (1). This contributes to substantial excess mortality for conditions that are amenable to health care, with 65% of excess mortality attributed to poor quality of care as of 2016 (2). This makes quality a key challenge for reaching the Sustainable Development Goals and a top priority of global health initiatives (3,4).

However, it is difficult to improve something that is not measured and a major obstacle to improving quality is the scarcity of data. Quality can be measured through supply-side availability (sometimes called structural quality), processes of care via direct observations, exit interviews and community surveys, audit or records, or community health outcomes (5). Facility surveys are sometimes conducted in one-off studies, especially randomized control trials; however, they rarely measure processes or health outcomes and the sampled nature of this data limits its usefulness in tracking progress over time (6). Previous proposals to improve monitoring have been made, yet little progress to date, especially in geographies with humanitarian crises where it is even harder to measure (7).

One source of data on quality of care is from the evaluations of performance-based financing (PBF) programs, which conduct pre-and post-intervention surveys in the same facilities and sometimes measure both processes and service coverage, which are rarely available outside of stand-alone vertical programs. Specifically, the World Bank has managed several PBF evaluations with a focus on primary health care, offering financial incentives to health facilities and sometimes health providers based on the quantity and quality of services (8). As a part of impact evaluations of PBF programs, detailed data was collected on processes and outputs of healthcare delivery through facility assessments linked to direct observations of clinical consultations, provider interviews, and exit interviews with healthcare users. In this analysis, we utilized one such dataset from the Health System Strengthening for Better Maternal and Child Health Results Project (PDSS) in the Democratic Republic of Congo (DRC) to examine the correlates of process quality for antenatal care (ANC) consultations (9).

This reanalysis of PDSS data adds to a growing body of research on the quality of care with a focus on ANC. Variations in quality have been documented between countries and across sub-national regions (10–12), however, to identify strategies for improving care, a deeper understanding of the reasons behind quality gaps are needed. Existing literature on antenatal care quality has examined a range of factors affecting its delivery and effectiveness, including the consistency of care, the availability of essential resources, adherence to clinical protocols by healthcare professionals, and the skill levels of providers, among others (1,10,13). Studies have found associations between the overall quality of ANC consultations to structural quality, follow-up training on ANC, and socioeconomic status of the pregnant individuals (14,15).

We built on this by examining which factors were associated with the completion of each individual prescribed action (e.g., administering the tetanus toxoid vaccine), rather than aggregating into an overall index, as has been done by others. This contrasts with the usual approach of associating explanatory factors with a composite index for process quality, typically calculated as the percentage of actions completed out of total assessed (10,12,14,16). This choice was driven by the hypothesis that while some factors may systematically affect the completion of all actions, others may be significant for only specific actions and thus could be overlooked in an aggregate analysis. Further, the disaggregated approach may reveal nuanced and potentially non-linear relationships that aggregate approaches might not be powered to detect. If these relationships do exist, there would be policy implications since the findings on action-specific factors can inform targeted interventions that may be more effective and less costly than system-wide investments.

In this study, we analyzed a dataset that combines direct observations of clinical care, exit interviews with users of ANC consultations, and facility assessments, to examine how supply-side readiness for each action affects its completion. Despite the established protocol for ANC consultations, recommended actions are not always done and we sought to understand the barriers to content of care completion. In theory, the completion of each recommended action may be affected by factors including but not limited to the facility’s availability of essential equipment and supplies (referred to as “essential items” hereafter), the provider’s knowledge of the recommended actions, the patient’s demand for specific actions, behavior patterns, and the sociocultural dynamics between the provider and the patient (12,14,15). We looked at how the completion rates differed by the type of care component (i.e., physical examinations, preventive treatments, or diagnostic tests), the training level and gender of the provider, the provider’s knowledge of ANC consultation, and the interaction between supply-side readiness and these other factors. We pinpointed specific areas for improvement and potential interventions to enhance the quality of ANC consultations.

## Methods

### Data

The data used in this study are from the impact evaluation of the World Bank-funded Health System Strengthening for Better Maternal and Child Health Results of Congo Project (PDSS) (17,18). As part of the project’s impact assessment, data was collected through health facility assessment surveys, interviews with health providers, direct observations of consultations (under-5 outpatient visits, ANC, and family planning), and patient exit interviews. Fifty-eight health zones from six provinces (Kwango, Kwilu, Mai-Ndombe, Haut Katanga, Haut Lomami, and Lualaba) of the DRC were included in both the baseline and endline surveys (19) and form the basis of this analysis. The surveys were conducted between June 2015 and March 2016 (baseline), and July 2021 and May 2022 (endline). During the baseline, five health centers and one referral hospital from each health zone were randomly selected to be surveyed. During the endline, the same facilities were re-visited or substituted if revisiting was not possible. Details of the randomization process and facility selection are described elsewhere (19).

In the baseline survey, the protocol was to conduct five observations of ANC visits at each of a subset of randomly selected facilities; in endline, up to eight observations were conducted in all participating facilities (15). This resulted in a total of 1,434 direct observations completed at baseline and 2,321 at endline. Of those, 406 (28%) in the baseline and 1,778 (77%) in the endline were first consultations (ANC1) with relevant facility-level readiness indicators available and were included in our study.

### First-visit ANC consultation

Our analyses focused exclusively on the first antenatal consultations, which are a useful analysis setting because the recommended procedures for ANC consultations are standardized and clearly associated with quality of care and health outcomes (10,20). Most recommended actions for ANC consultations should be performed for all pregnancies (e.g., blood pressure should be taken and a urine sample should be analyzed during all ANC consultations to screen for signs of preeclampsia) (20,21). Further, by restricting to the first visits only, we minimized the risk of appropriate variations in content of care that could be caused by visit history and progression of the pregnancy, which could otherwise confound the analysis.

In this analysis, we examined how contributing factors from different sources (i.e., the facility, the provider, and the patient) were associated with the completion of the actions and the relative importance of each contributing factor.

### Outcome variables

Using the survey questionnaire for direct observations of ANC consultations, we compiled a list of questions documenting whether the interviewer observed the completion of each of the recommended actions during the ANC consultations. We followed Fink et al. (15) and started with 20 recommended actions for the first ANC visit and classified them by component (Table 1). We used the completion of each of these twenty individual actions as a primary outcome variable in our analyses. See Table S1 in Additional file 1 for survey question details.

**Table 1.** List of actions expected to be completed during ANC1 and the required facility inputs.

| Component | Recommended action | Required facility inputs |
| --- | --- | --- |
| Physical examination | Take the weight of the pregnant individual | Adult scale |
|  | Take the pregnant individual's blood pressure | Blood pressure cuff, stethoscope |
|  | Measure the fundal height | Tape measure |
|  | Perform chest auscultation | Stethoscope |
|  | Listen to the fetal heartbeat | Obstetric stethoscope |
| Diagnostic tests | Perform a urine test for albumin and glucose | Urine analysis kit |
|  | Offer or refer the pregnant individual for voluntary HIV testing | HIV test kit |
|  | Offer or refer the pregnant individual for a syphilis test | Test for syphilis |
|  | Ask for a blood sample and measure the hemoglobin level | Hemoglobinometer |
| Preventive treatments | Prescribe or give a tetanus toxoid shot | Tetanus Vaccine |
|  | Prescribe or give iron supplements | Iron (with or without folic acid) |
|  | Prescribe or given intermittent preventive treatment of malaria | Sulfadoxine-Pyrimethamine (SP) |
|  | Give the pregnant individual a treated mosquito net | Insecticide-treated net |
| Screening for danger signs | Ask about the HIV status of the pregnant individual. | Doesn't require any equipment or supplies. |
|  | Ask for the Rh factor of the pregnant individual. |  |
|  | Ask about the medications the pregnant individual is taking |  |
|  | Ask about tetanus vaccination status of the pregnant individual. |  |
|  | Ask about symptoms experienced during the current pregnancy |  |
| Counseling | Give advice on nutrition |  |
|  | Give advice on danger signs and risk factors during pregnancy |  |

Table 1: Component categories and required facility inputs were assigned by the authors with expert input. All recommended actions for ANC1 that were observed were included in the analysis. HIV = human immunodeficiency virus. Rh = Rhesus factor D antigen.

Our analyses diverged from the conventional approach in that we treated the observed completion status of each recommended action during the first ANC consultation as the primary outcome variable. Doing so allowed us to assess the relationship between specific contributing factors and each individual action, in order to check for dynamics that may not be detectable when the outcome is aggregated.

### Independent variables

We constructed independent variables to capture factors that could impact the completion of a recommended action, corresponding to health facility attributes, health provider knowledge, health provider attributes, and patient attributes.

Health facility attributes included facility location, facility type, province, and availability of essential items (Table S1, S2 in Additional file 1). The availability of an essential item was coded as binary indicating available or not, based on the reported quantity of functional equipment and medicines, as well as the number of days that medicines were out of stock. Equipment, medicines, and testing kits were considered available if the quantity reported as functional and available was greater than zero. As a sensitivity test, we also considered the essential item to be available if the medicine was out of stock on the day of facility assessment but had fewer than 15 days of stockout in the previous 30 days. Using this alternative definition, we found the results were not substantially impacted (results not shown).

The health provider attributes included gender and qualification. We used Section 1 of the direct observations survey of ANC consultations to construct the variables. The gender and qualification composition of the providers of first ANC consultations were summarized in Table

S2 in Additional file 1. In the survey, the nurses can be further differentiated into A3, A2, L2 and A1 based on level of training (22–24). Nurses at levels A3 and A2 received training at secondary-school level, while nurses at levels L2 and A1 received training at higher-education level equivalent to a bachelor’s degree (23,24).

The health providers’ knowledge variable was constructed using responses from the vignette on the provision of ANC consultation found in Section 11 of the provider interviews survey. The providers were given a list of care options, and they were asked to identify the questions that they should ask the pregnant individual and the actions that they should take during a first ANC visit. We coded their response “1” if they responded that the action should be performed, and “0” if not (Table S1 in Additional file 1).

The patient attributes were retrieved from the exit interviews survey of ANC consultations. The distributions of age and educational attainment of pregnant individuals in exit interviews of first-visit ANC consultations were summarized in Table S2 in Additional file 1. We constructed a wealth index following Fink et al (15) (Table S3 in Additional file 1). Other variables related to the patient included an indicator for whether this was the individual’s first pregnancy and the gestation age in weeks, as reported in Section 1 of the direct observations survey of ANC consultations (summary of variables in Table S2 in Additional file 1).

### Level of analysis

The unit of observation in our analyses was an individual consultation, with each recommended action examined separately. We examined baseline and endline observations separately in order to see if the determinants of quality of care changed during the course of the intervention.

### Bivariate analyses of equipped facilities

Action completion, availability of essential items, and provider knowledge were summarized by frequency and proportion. To assess whether changes in these variables were statistically significant between baseline and endline, we conducted chi-squared proportion tests.

To assess whether the availability of the essential item significantly predicted the likelihood of completing the action, we calculated the completion rate separately for observations where the essential item was available and where it was not. Chi-squared tests were used to compare the two proportions. Some facilities reported that they did not have a room for storing medicines and supplies, which caused a skip pattern in the survey and resulted in missing data; care observations from these facilities were excluded from this part of the analysis.

To investigate the influence of provider characteristics on action completion, we stratified observations by provider gender and qualification and categorized the providers into four groups based on relevant qualifications: ‘midwives’, ‘A3 and below’, ‘A2’, and ‘A1/L2/MD’. We cross-checked qualifications with years of education to verify the appropriateness of the groupings.

We then conducted proportion tests on the completion rate of each group versus the highest completion rate observed among all groups. Thus, the null hypothesis was that completion rates observed across groups of providers were the same regardless of level of training or gender.

We examined the effect of training by conducting proportion tests among female providers across different training levels. We examined the effect of provider gender by comparing completion rates between female providers and male providers at the ‘A2’ and ‘A1/L2/MD’ training levels, respectively.

For this part of the analysis, we examined only observations where the essential item was available to eliminate the risk of confounding due to providers with higher training levels working at better-equipped facilities. Additionally, we limited the analysis to subgroups with at least 10 observations.

### Subgroup analyses on limiting factors

To explore the potential causes of unperformed actions, we categorized incompletions based on the availability of the essential item at the facility and the provider’s knowledge of recommended actions. For each recommended action, we decomposed incompletions using the three-gap model (11,25) into the following 4 types: facility readiness gap, provider knowledge gap, know-do gap, and dual deficiency. Table 2 illustrates each scenario using syphilis testing as an example. Additionally, we calculated the decomposition separately for providers of different levels of training, to compare how the decomposition differs among providers with varying levels of training. We did not calculate or report decomposition when the total number of observations in the segment was fewer than 10.

**Table 2.** Descriptions for categories of incompletion using syphilis testing as an example.

| Category | Description for one example action |
| --- | --- |
| Facility readiness gap | Syphilis testing kits <u>were not available</u> at the facility, and it was not completed. |
| Provider knowledge gap | The provider <u>did not know</u> a syphilis test should be conducted during the consultation, and it was not completed. |
| Know-do gap | The facility <u>had</u> syphilis testing kits, and the provider <u>knew</u> a syphilis test should be conducted during the consultation, but it was not completed. |
| Dual deficiency | The facility <u>did not have</u> syphilis testing kits, AND the provider <u>did not know</u> a syphilis test should be conducted during the consultation, and it was not completed. |

Table 2: Categories of incompletions were created based on the presence of one, both, or neither of the two limiting factors: the availability of the essential item at the facility and the provider’s knowledge of recommended actions.

### Cross-sectional regressions

To estimate the impact of the essential item’s availability on the completion of a recommended action, we fitted logistic regression models to analyze the probability of completion while controlling for additional sources of variation. The facility-level control variable was facility location. The provider controls included qualification and gender. The patient controls included age, educational attainment, and wealth quintile of the pregnant individual, the age of gestation (trimester), and previous pregnancy. We fitted a separate model for each action and used backward stepwise selection to choose the best-fit parsimonious model.

For the regression analyses, we examined 13 actions from three care components: physical examinations, diagnostic tests, and preventive treatments. These actions required at least one essential item. For example, measuring the pregnant individual’s blood pressure was dependent on the availability of a blood pressure monitor and a stethoscope (21), so these indicators were used for that outcome measure.

Since we ran 13 regressions on the same set of predictors, we faced increased risk of any predictor testing significant (p<0.05) just by chance, so we applied Bonferroni corrections (26).

For bivariate analyses, we reported the results from baseline and endline data separately. For cross-sectional regressions, we reported only the results from the baseline data to focus on the multivariate relationships before the PBF interventions. The statistical significance of the proportion tests and the regression results were reported in Additional file 1.

## Results

### Descriptive statistics on completion, readiness, and knowledge

The baseline sample contained direct observations of 406 first-visit ANC consultations conducted by 181 health providers at 167 health facilities. The endline sample contained direct observations of 1,778 first-visit ANC consultations conducted by 391 health providers at 329 health facilities. ANC consultations were almost universally performed by nurses or midwives (98% baseline, 99% endline). Most consultations were conducted by female providers (72% baseline, 84% endline). Only the health providers present on the day of provider interviews completed the vignette on the provision of ANC consultation, resulting in knowledge assessments for 128 providers in the baseline and 80 providers at the endline.

Recommended actions were not always completed during visits. At baseline, half of the actions were completed less than 50% of the time. Among the five components of ANC consultations, physical examinations had the highest rates of completion, and diagnostic tests had the lowest rates of completion. There was a statistically significant increase (p<0.05) in completion rate for 6 of the 20 actions, with improvement ranging from 8.8% to 38.2% (Table S4 in Additional file 1). Preventive treatments improved the most and physical examinations the least (Figure 1).

**Figure 1.**
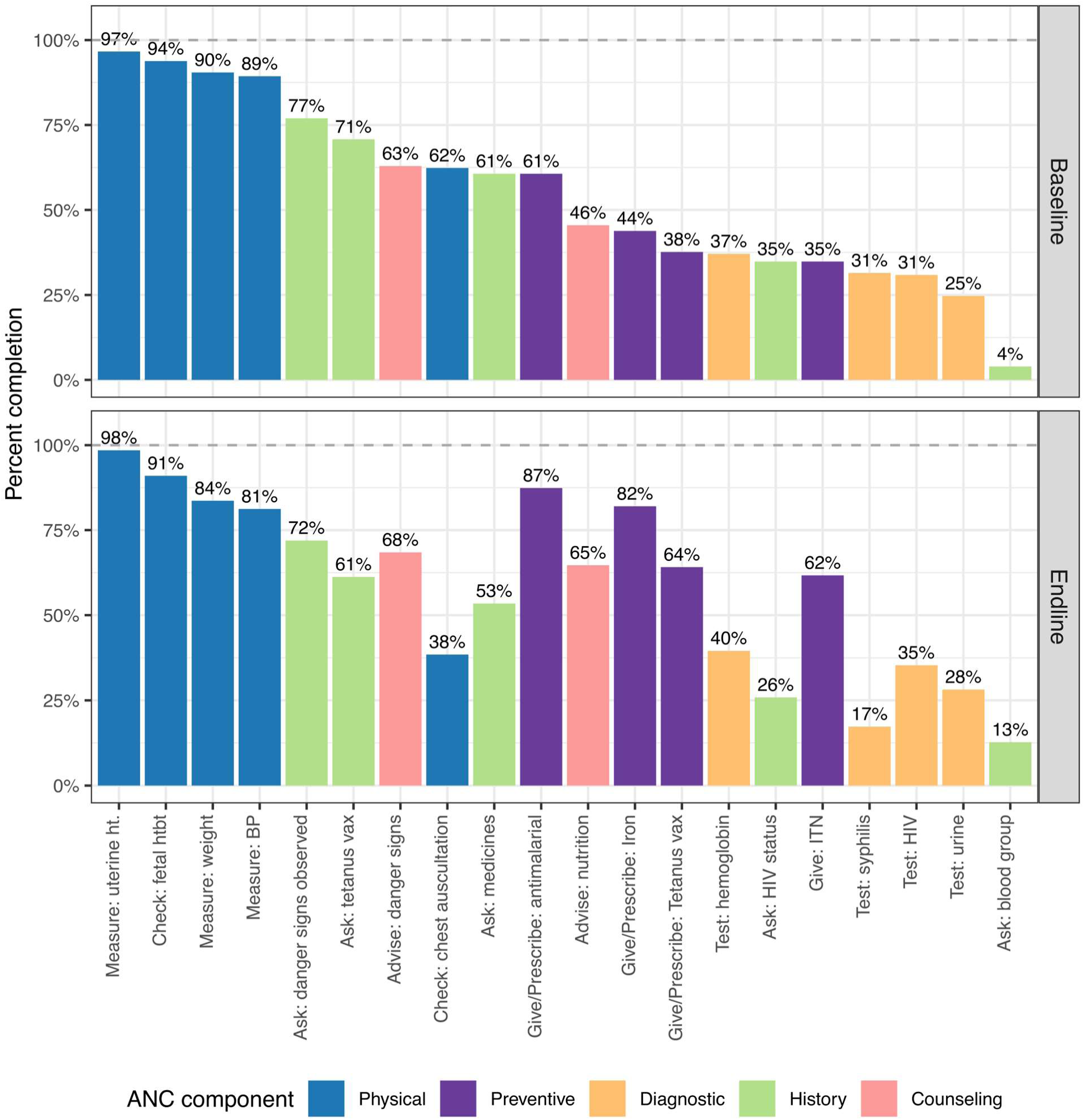
Completion rate of recommended actions during ANC1 consultations. Height of the bar indicates the percentage of all first antenatal care visits that were observed where a particular action was completed. All actions are recommended by WHO guidelines for the first visit. ht = height. htbt = heartbeat. BP = blood pressure. vax = vaccination. HIV = human immunodeficiency virus. ITN = insecticide treated net. ANC = antenatal care. Baseline survey was conducted in 2015-16. Endline survey was conducted in 2021-22.

Each of the physical examinations, diagnostic tests, and preventive treatments required some essential item (i.e., testing kit, medicine, or other supply) and availability of these items increased between baseline and endline. In the baseline, physical examinations had the best availability of essential items; all were above 80% and measuring tape and stethoscope exceeded 90%. In contrast, diagnostic tests were commonly unavailable, ranging from a high of 62% for the hemoglobin test to 14% for the urine test (Figure 2). There was a statistically significant increase (p<0.05) in the availability of seven of the thirteen essential items, with medicines and supplies for preventive treatments improving the most (Table S5 in Additional file 1).

**Figure 2.**
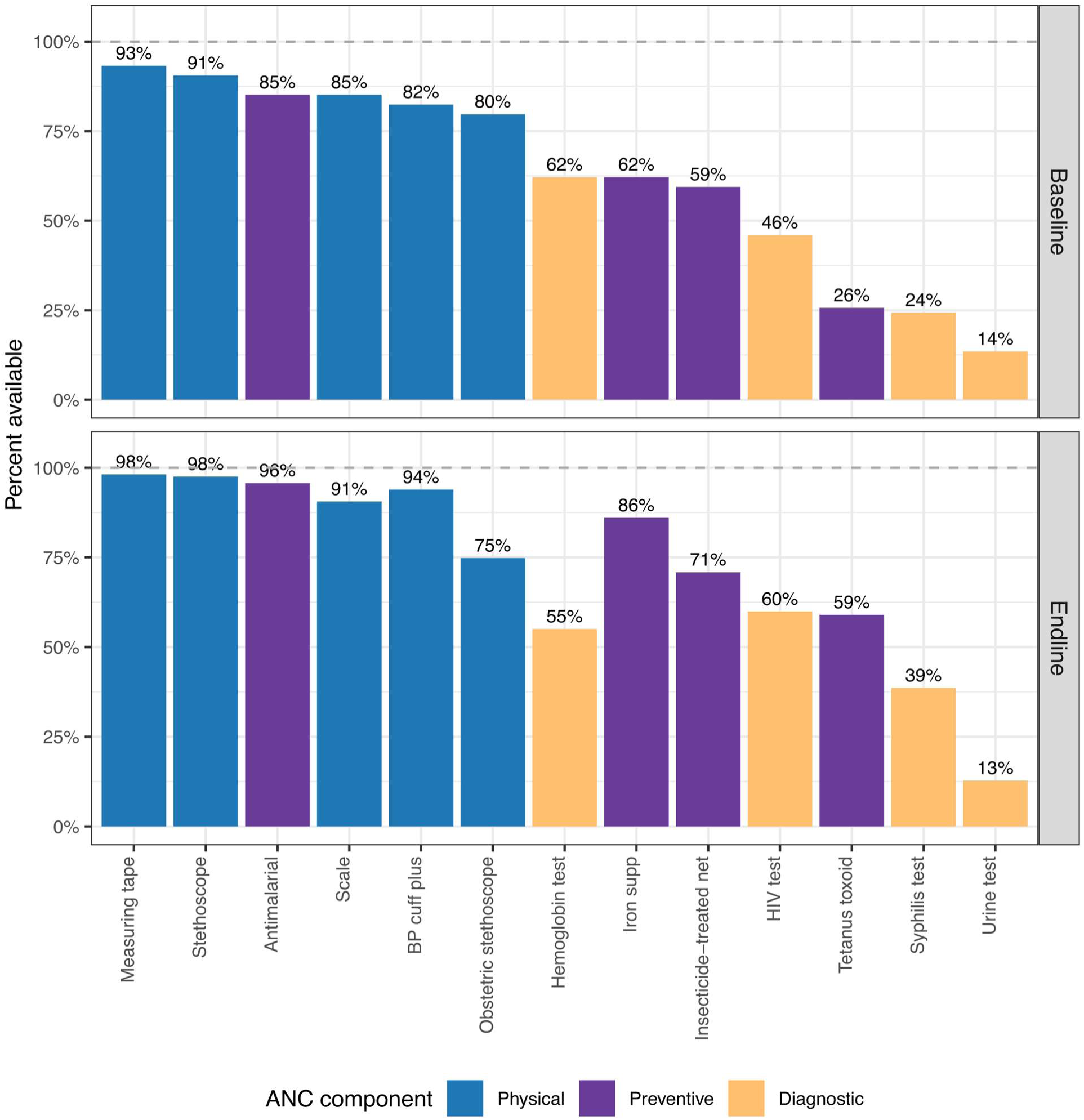
Availability of essential items at facilities. Height of the bar indicates the percentage of facilities where an essential item was available on the day of the survey. BP cuff plus = blood pressure cuff and stethoscope. Iron supp = iron supplement. HIV = human immunodeficiency virus. Baseline survey was conducted in 2015-16. Endline survey was conducted in 2021-22.

### Impact of facility readiness on completion of actions

We compared the completion rates when the essential item was available versus unavailable using chi-squared tests (Table S6 in Additional file 1). The tests were statistically significant (p < 0.05) for most of the thirteen actions, ten at baseline and eleven at endline. Differences were all positive, indicating that patients visiting better-equipped facilities were more likely to receive the relevant recommended action.

However, availability of the essential item did not fully explain variance in completion rates. First, we observed that even when the facility had the essential item, the action was not always completed. For example, diagnostic tests were performed less than half of the time (range 32-49% in the baseline and 24-55% in the endline) even when the testing kit was available (i.e., the kit was on the shelf, but was not used during the consultation). Further, actions were sometimes observed as completed, even in facilities where the essential item was unavailable at the time of the survey. This was most common for physical exams (range 27-97% in the baseline and 8- 100% in the endline) and least for diagnostics (range 13-30% in both endline and baseline) (Figure 3).

**Figure 3.**
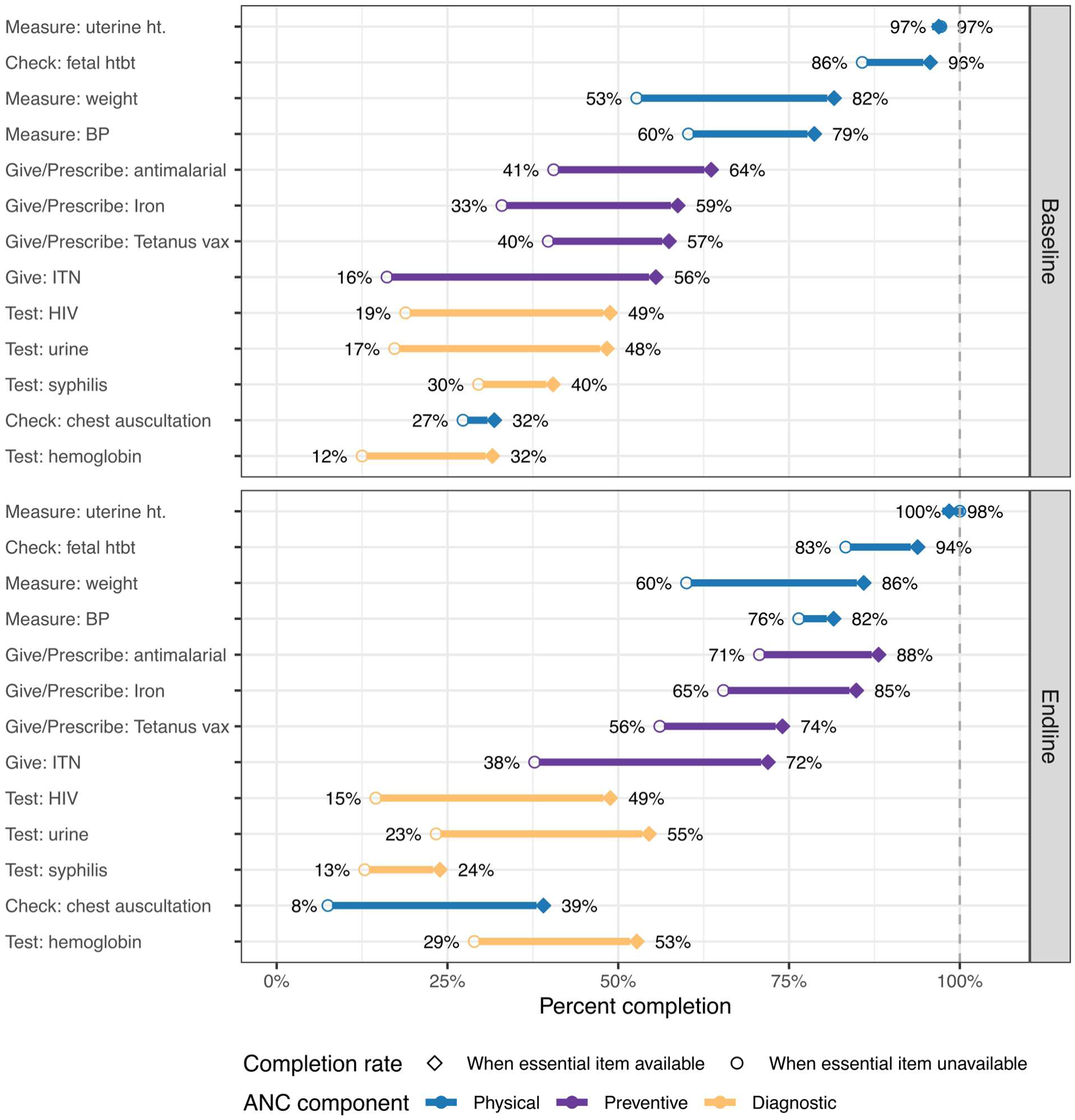
Completion rate of actions, conditional on essential item availability. Percentages indicate the proportion of all first antenatal care visits that were observed where a particular action was completed. Hollow circle indicates completion rate when the essential item was not available. Filled diamond indicates the completion rate when the essential item was available. Solid line indicates the difference between. All actions are recommended by WHO guidelines for the first visit. ht = height. htbt = heartbeat. BP = blood pressure. vax = vaccination. HIV = human immunodeficiency virus. ITN = insecticide treated net. ANC = antenatal care. Baseline survey was conducted in 2015-16. Endline survey was conducted in 2021-22.

### Impact of provider characteristics on completion of actions

We calculated the completion rate for actions by provider training level and gender, and found meaningful differences for some activities. Inter-group differences were small for physical examination actions and rarely statistically significant, whereas larger and significant disparities were observed in both preventive treatments and diagnostic tests. (All calculated percentages were listed in Table S6 in Additional file 1.) In the baseline, highly-trained female providers had the highest completion percentages for eight out of thirteen actions and were not statistically different from the best performing group for the other five. Among female providers, higher training levels were generally associated with better completion rates for both preventive treatments and diagnostic tests. For example, for HIV testing, rates increased from 25% to 32%, 58% and 93% for providers with additional training (Figure 4).

**Figure 4.**
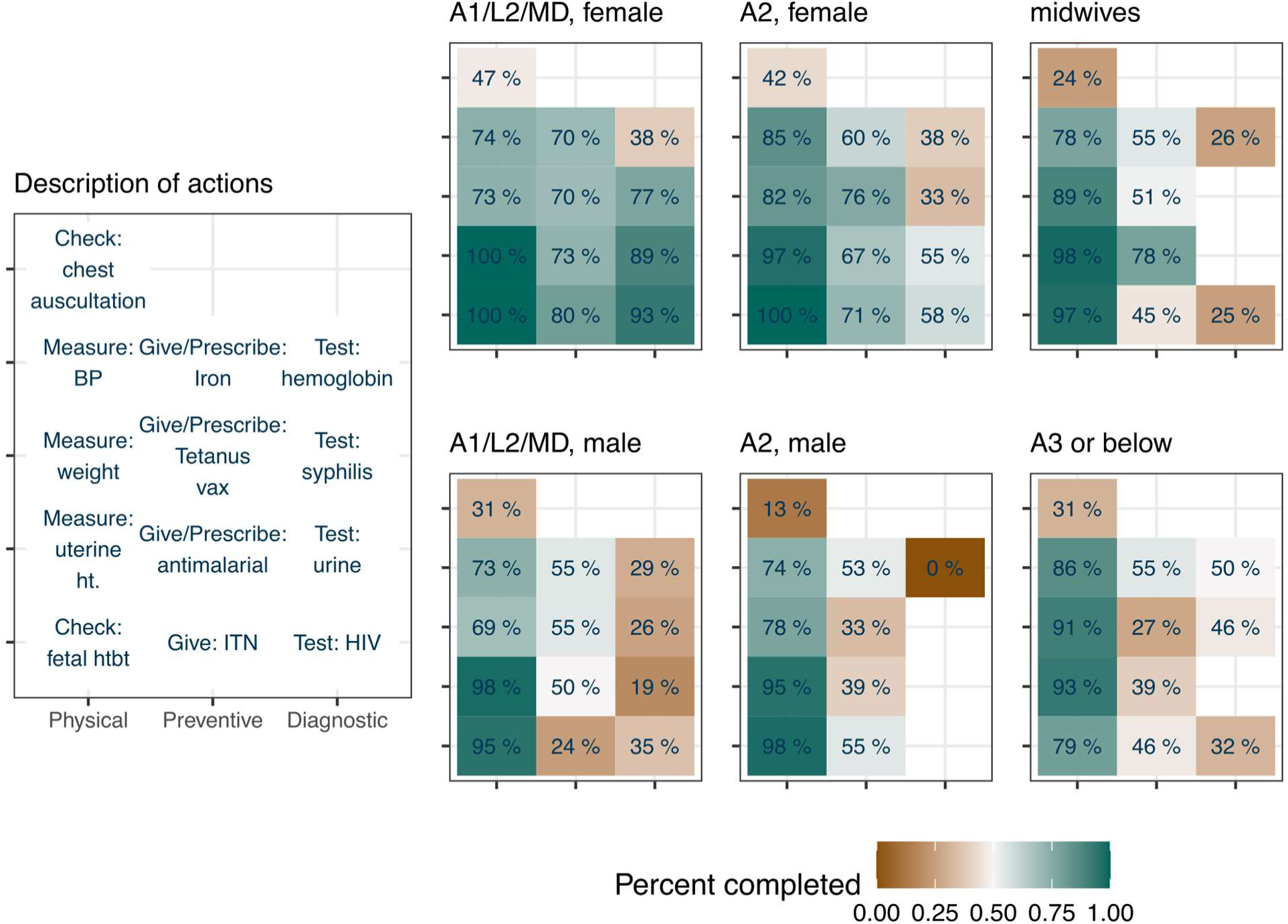
Completion rate by provider characteristics. Percentages indicate the proportion of visits for which a particular action was completed during a first ANC (ANC1) visit. Blank cells indicate that there was inadequate sample size (fewer than 10 visits) to calculate the percentage. Headers indicate the level of training and the gender of the provider who conducted the visit. There were thirteen recommended actions that required an essential item, each of which was described in the reference panel and detailed in Table S1 in Additional file 1. A1/L2/MD = Medical doctors and nurses with L2 and A1 training. A2 = nurses with A2 training. A3 = nurses with A3 training. BP = blood pressure. Vax = vaccination. Ht = height. ITN = insecticide treated net. HIV = human immunodeficiency virus. Calculations used baseline survey conducted in 2015-16.

Comparing the completion rates of male and female providers with equal training, we found that male providers underperformed female providers, particularly in completion of diagnostics and preventative treatments, as well as specifically chest auscultation (listening to the respiratory system, ideally with a stethoscope) (Figure 4). For example, among nurses of level A2, female providers were 3.2 times as likely to perform chest auscultations, and 2.3 times as likely to prescribe tetanus vaccination as their male counterparts. Among the highest-trained providers (“A1/L2/MD”), female providers were 2.7 times as likely to give HIV tests, 4.7 times as likely to give urine tests, and 3.0 times as likely to give syphilis tests as male providers with equivalent training.

### Limiting factors for the completion of actions

For visits when an action was not completed, we categorized these incompletions in the context of limiting factors as: facility readiness gap, provider knowledge gap, know-do gap, and dual deficiency, to examine variations in the likely causes for unperformed actions. We present the results by care components and provider training levels (Figure 5).

**Figure 5.**
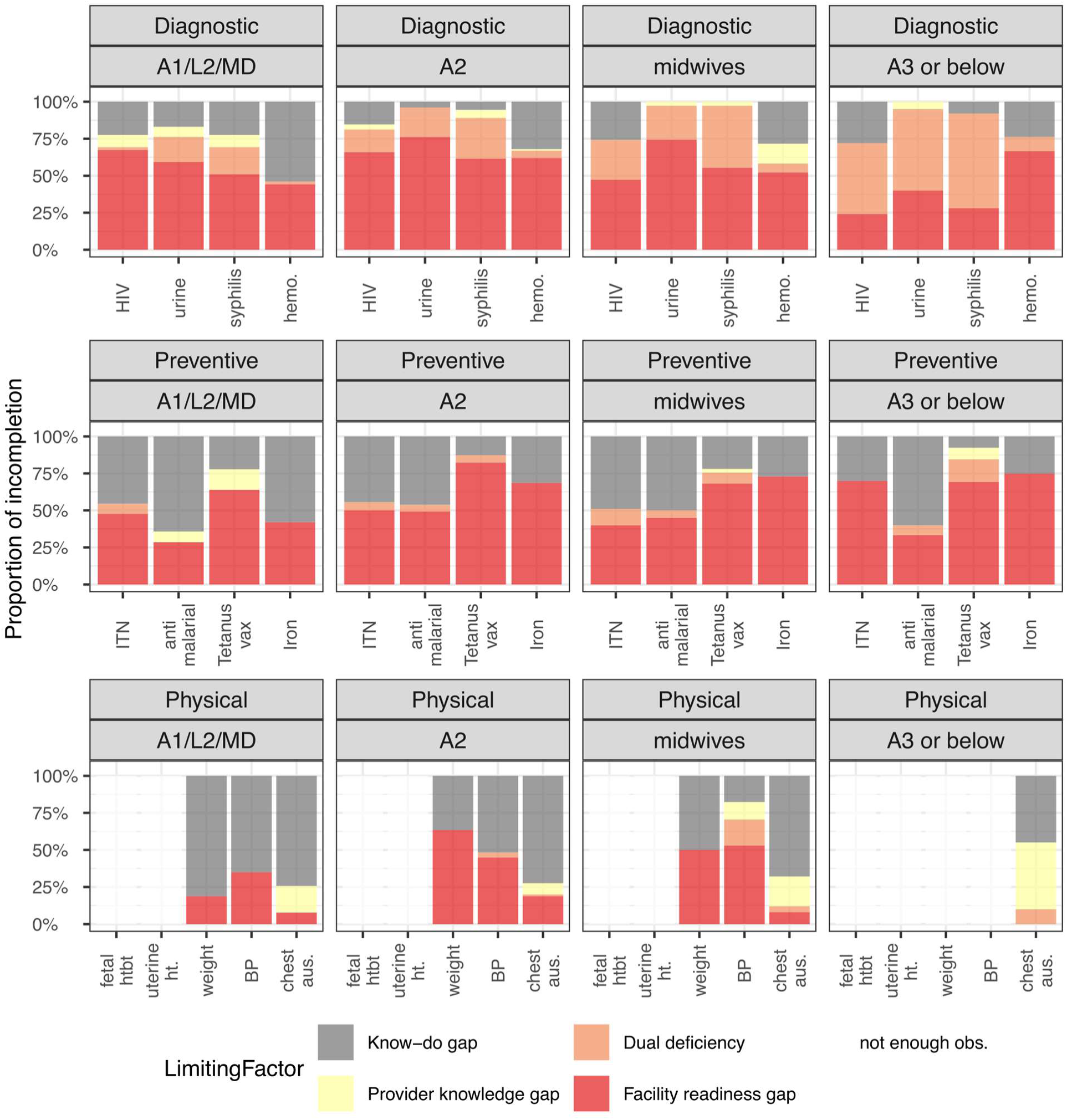
Factors limiting action completion, by provider training level. Bar height indicates the proportion of antenatal care visits that fell into one of four groups: facility readiness (provider knew to do the task, but essential item not available), provider knowledge (essential item available, but provider did not know to do the action), dual deficiency (both readiness and knowledge gaps), or know-do gap (neither readiness nor knowledge gap). A1/L2/MD = Medical doctors and nurses with L2 and A1 training. A2 = nurses with A2 training. A3 = nurses with A3 training. HIV = human immunodeficiency virus. Hemo. = hemoglobin. ITN = insecticide treated net. Vax = vaccination. Htbt = heartbeat. Ht = height. BP = blood pressure. Aus. = auscultation. Calculations used baseline survey conducted in 2015-16.

In the baseline sample, stockouts of testing kits were the most frequent limiting factor for diagnostic testing, contributing to 79% of all incompletions across all provider levels.

Approximately two-thirds of these were attributable to stockouts only, and the remaining one-third of visits had a dual deficiency. Generally, providers with less training had a higher rate of dual deficiency (10% among “A1/L2/MD” versus 44% among “A3 or below”), indicating that providers with lower level of training had less knowledge of diagnostics.

Stockouts of medicines and supplies were also the most frequent limiting factor for preventive treatments in the baseline sample, accounting for 60% of all incompletions. This changed in the endline, declining to 29% of all incompletions due to significant improvements in availability of preventive treatment medicines and supplies. A further 37% of incompletions in the baseline (40% in the endline) were due to the know-do gap and were not caused by any apparent supply-side constraint.

Physical exams where the care component completed most consistently, with the know-do gap being the largest contributor to their incompletion, at 54% in the baseline sample (28% in the endline) across all actions and provider types. Generally, the know-do gap was larger for better-trained providers, indicating that there may be other barriers to care that were not directly observable in this dataset.

Across the three care components, we found that both stockouts and provider’s knowledge hindered completion of actions among providers with lower training levels, while the know-do gap was more prevalent among providers with higher training levels.

### Results from cross-sectional regressions

We conducted cross-sectional regression analyses using the baseline sample to estimate the impact of availability of the essential item on the completion of each recommended action, while controlling for potential confounding factors. We selected a best-fit parsimonious model for each action and summarized the results from 13 regressions in Figure 6. The pseudo R-squared ranged from 0.07 for explaining the completion of measuring blood pressure, to 0.41 for explaining the variation in completion of urine tests.

**Figure 6.**
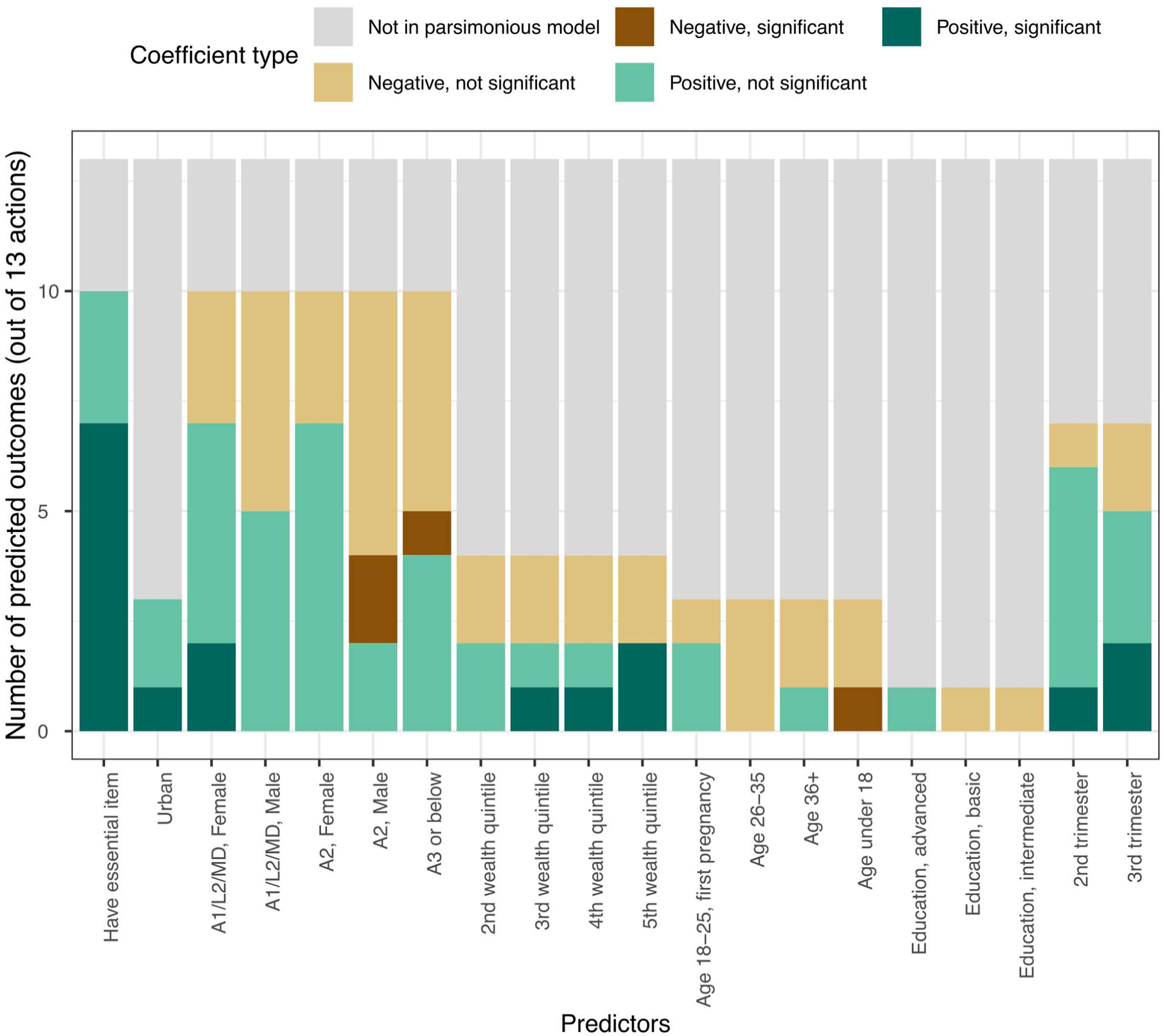
Summary of significance for regression variables. Bar height indicates the number of regressions for which a variable was significant or not at the p <0.05 level after applying the Bonferroni correction. The maximum possible height is thirteen, one for each regression that was run. The reported results are relative to a baseline consultation taking place at a rural facility without the essential item, provided by a midwife, where the pregnant individual was aged between 18 and 25 with no formal education, had a previous pregnancy, and was visiting during the first trimester.

As a validity check, we checked the results for regressions on the completion of measuring uterine heights and checking fetal heartbeat, since these should primarily happen in later stages of pregnancy and found that the estimated coefficient for age of gestation (trimester) was positive and statistically significant for completion of both actions, aligned with appropriate clinical practice.

Availability of the essential item was the most important predictor overall, being positively associated with completion of all actions, and statistically significant (p<0.05) for 7 out of 13 actions, most notably for preventive treatments. For example, after controlling for other covariates, the odds of action completion were 5.9 [CI: 3.4, 10] times higher for giving the patient an insecticide-treated net (ITN) when the supply was available at the time of the facility survey, and 3.1 [CI: 1.7, 5.7] times higher for giving or prescribing iron when the medicine was in-stock at the time of the facility survey. Estimated coefficients and associated p-values can be found in Table S7 in Additional file 1.

Provider attributes were the next most frequent significant predictors for completion: including provider attributes improved model fit for predicting 10 out of 13 actions, consistent with our findings in the bivariate analyses. Among female healthcare providers, completion was generally positively associated with training level and most evident in diagnostic tests. For example, compared to midwives, the odds of prescribing an HIV test by female A2 nurses were 2.9 [CI: 1.3, 6.4] times higher, and 8.0 [CI: 3.1, 20.7] times higher for female providers in the group “A1/L2/MD”, after controlling for other covariates. The estimated odds of completion were consistently higher for male providers in group “A1/L2/MD” compared to male A2 nurses across actions. We note that male providers conducted 26% of the observed consultations, leading to a smaller sample size compared to female providers and thus weaker statistical power.

Patient attributes were infrequently significant in predicting the completion of activities, and for some vulnerable groups, their demographics were negatively associated with outcomes. Patient age was included in the final model for predicting three actions, and patient educational attainment was significant for only one action. The age and pregnancy history of the pregnant individual were not significant in predicting completion of most actions, however, the age group of 18 and younger stood out for being negatively associated with completion of three actions, despite a small sample size (N=26, 7.9% of total observations). Comparing a patient under the age of 18 to a patient aged between 18 and 25 with previous pregnancy, the odds of receiving tetanus vaccination were 25 times higher for the older group (OR = 0.04 [CI: 0.004, 0.34]) and the odds of receiving an ITN were 5 times higher for the older group (OR = 0.20 [CI: 0.06, 0.61]). The patient’s educational attainment was not a significant predictor for completion of most actions; higher educational attainment was not associated with increased likelihood of completion except for the highest level of education in predicting completion of hemoglobin test.

## Discussion

Methodologically, we found that it was valuable to evaluate structural quality at a more detailed level than is usually done, considering the requirements needed to deliver individual components of care. More specifically, using a typical composite index to measure structural quality risks obscuring poor availability of some medicines essential for pregnancy-related preventive treatments (e.g., iron supplements) by averaging in the better availability of general equipment for physical exams. This approach may miss opportunities for targeted investments to improve quality of care.

The positive and significant association between the availability of essential items and the completion of recommended actions indicated that low structural quality was a critical constraint for providing high quality of care, especially for diagnostic tests and preventive treatments. This suggested that future investments in structural quality could yield larger marginal impact when directed at these two care components, compared to physical exams where the provider know-do gap was a bigger contributor to the remaining quality gap.

However, taken together, the analyses showed that supply-side factors were not the only barriers to quality of care. First, the completion rates were substantially below 100%, even when the essential items were available, which showed that structural quality was a necessary but not sufficient condition and alone did not guarantee high quality ANC. Further, while professional training was associated with higher completion rate among female providers for diagnostic tests, this was not true for physical exams or preventive treatments and the know-do gap remained to some extent for all actions.

Since diagnostic tests are more complex than physical exams or preventive treatments, the impact of training level on completion of diagnostics points to two potential interventions: either ensuring that higher-trained providers take responsibility for this aspect of ANC or upskilling lower-level providers to ensure they have the knowledge required to complete diagnostics.

Taken together with the difference in performance between male and female healthcare providers of equal training, the remaining know-do gap indicated that care provision depended on more than supply-side availability. Cultural barriers likely also existed and in order to address these: a) care delivery should be designed to align with community expectations and values, and b) the community should be educated on what to expect and how to effectively engage with providers to ensure all actions are completed safely. In addition, provider behavior and compliance with standard care protocols needs to be supervised and reinforced in order to address any issues with follow-through.

Although previous literature has suggested that provider effort is a key contributor to variations in completing actions (25), with the available dataset, we could not directly assess whether there was a difference in provider effort or if socio-cultural barriers were instead the larger challenge. Given that we observed the largest differences in actions to be those that could be perceived as sensitive, such as physical exams requiring bodily contact or diagnostic tests involving handling blood or urine, we suspect that interpersonal dynamics are playing an important role, but this needs further investigation to be confirmed. Of note, evidence showed a difference in care provision between male and female providers, as well as generally low performance for tasks requiring close patient contact, such as chest auscultation—even among highly trained female providers.

These dynamics warrant further investigation into whether this is driven by provider hesitancy, resistance from the pregnant person, or other sources, in order to understand how to create a safe environment for the provision of high-quality care. Depending on findings, feasible approaches may include provider training, patient education, or new technologies that support more comfortable interactions, with the choice of approach guided by the results. Additionally, data collection efforts with an emphasis on documenting provider-patient interactions in varied cultural contexts could facilitate such investigations.

The decomposition of incompletions confirmed that improvement in availability of supplies alone would not fix all incompletions. Both unavailability of essential items and providers’ lack of knowledge on recommended actions contributed to incompletions, but the impact of each limiting factor varied for different care components. Stockouts were present in more than half of the incompletions for diagnostic tests and preventive treatments, but they were much less common for physical exams. Additionally, diagnostic tests had the highest proportion of incompletions due to concurrent stockouts and providers’ lack of knowledge, which indicated that interventions need to target both gaps in order to impact health outcomes. The decomposition showed that an improvement in medicine availability should increase preventive treatments, but it would be inadequate for diagnostics. Thus, interventions need to pair improved availability of testing kits with complementary training on protocols, particularly for providers with a less comprehensive degree program.

The decomposition analysis also showed that know-do gaps were a challenge across all care components, most notably for physical examinations and among providers with more advanced training. This was expected, since know-do gaps are revealed only after constraints on structural quality and providers’ knowledge have both been eliminated. However, when the know-do gap is the main contributor to a persistently low completion rate, like in the case of chest auscultation, it is important to understand what socio-cultural barriers underlie the remaining gap so that appropriate interventions can be planned.

We observed a notable proportion of action completions for all care components even when the necessary equipment or medical supplies were unavailable. For physical examinations, one plausible explanation was that alternative methods were used when the required equipment was unavailable. For example, the uterine height might have been measured through visual inspection and the fetal heartbeat measured by hand. This phenomenon was less frequent but also present in preventive treatments and diagnostic tests, and we suspected two potential causes. Firstly, the availability of essential items was assessed during facility evaluations, which occurred at different times than the consultations, meaning that stock levels could have changed in the interim. Additionally, since the survey questionnaire did not distinguish giving a medicine or test from prescribing a medicine or test, it is possible that when the action was observed to be completed when the medicine was unavailable, the provider wrote a prescription for the item rather than dispensing the medicine to the patient.

We could not test these hypotheses with currently available data, limited by the binary responses to the survey questions regarding completion, but anecdotal experience reported to us by providers from similar settings supported these hypotheses. One concern is that these alternative methods may be less effective than the recommended protocols and may lead to delays or errors in diagnosis, thus resulting in lower quality of care despite the high completion rate. This is an area where data collection with an emphasis on both qualitative and quantitative measures of the process of care would greatly help.

This study has several limitations. First, in predicting completion of individual actions we included selective predictors that we deemed most relevant during the care process, but this approach may have exposed us to the risk of omitting important variables. Second, stratifying the sample by provider characteristics and the availability of essential items may lead to the creation of subgroups with very few observations, which can result in estimates that have large standard errors. We encountered similar challenges in assessing the difference in completion of actions between female and male providers of equal training. Consequently, while some estimated coefficients were consistent with the hypothesized relationships, they were not statistically significant, suggesting that we may not have detected a true relationship. Third, we were unable to empirically examine the reasons for differences in performance between female and male providers due to a lack of appropriate measures for cultural contexts.

## Conclusions

This study added to existing evidence on factors affecting the quality of ANC consultations. Supply-side intervention could improve service quality for ANC1 visits. This was especially true for improved product availability for preventive care (e.g., vaccination) and for provider qualification. However, availability and training alone were insufficient to ensure the completion of all recommended actions and the know-do gap could remain, which we observed for physical examination actions (e.g., chest auscultation). Combined with the quality gap between female and male providers, this suggested that there are additional cultural barriers to achieving the highest possible quality of care. Thus, we concluded that while supply-side interventions are necessary and impactful, policy makers also need to invest in understanding and then addressing the patient-provider dynamics of an ANC1 visit, in order to achieve the best health outcomes for all.

## List of abbreviations

ANC: antenatal care
ANC1: first antenatal consultation
BP: blood pressure
DRC: Democratic Republic of Congo
HIV: human immunodeficiency virus
ITN: insecticide-treated net
LMIC: low-and middle-income countries
OR: odds ratio
PBF: performance-based financing
PDSS: Projet de Développement du Système de Santé

## Data Availability

All data used in the analysis that is described in this manuscript comes from the World Bank microdata library. (https://microdata.worldbank.org/) The baseline survey can be referenced by ID: COD_2015_HRBFIE-FBL_v01_M and the endline survey can be referenced by ID: COD_2021-2022_HRBFIE-HFFU_v01_M.

## Ethics approval

Not applicable.

## Consent for publication

Not applicable.

## Author contributions

BH and RH conceptualized the study and drafted the manuscript. RH conducted data cleaning, analysis, and developed the visuals. BH supervised the analysis, provided critical feedback, and edited the manuscript. Both authors reviewed and approved the final version of the manuscript.

## Acknowledgements

The authors would like to thank Gil Shapira and Supriya Madhavan for providing data access and their support and insight into interpretation of the results from this study.

## Additional material

Additional file 1.xlsx contains supplementary tables providing details for variable definitions, statistical significance of proportion tests, and regression results.

